# Development and utilization of an intelligent application for aiding COVID-19 diagnosis

**DOI:** 10.1101/2020.03.18.20035816

**Authors:** Zirui Meng, Minjin Wang, Huan Song, Shuo Guo, Yanbing Zhou, Weimin Li, Yongzhao Zhou, Mengjiao Li, Xingbo Song, Yi Zhou, Qingfeng Li, Xiaojun Lu, Binwu Ying

**Author notes:** Zirui Meng, Minjin Wang, Huan Song. contributed equally to this work. Correspondence author: Binwu Ying. No.37 Guoxue Alley, West China Hospital, Sichuan University, Chengdu 610041, Sichuan Province, P. R China.

## Abstract

**Background:** COVID-19 has been spreading globally since emergence, but the diagnostic resources are relatively insufficient.

**Results:** In order to effectively relieve the resource deficiency of diagnosing COVID-19, we developed a machine learning-based diagnosis model on basis of laboratory examinations indicators from a total of 620 samples, and subsequently implemented it as a *COVID-19 diagnosis aid APP* to facilitate promotion.

**Conclusions:** External validation showed satisfiable model prediction performance (i.e., the positive predictive value and negative predictive value was 86.35% and 84.62%, respectively), which guarantees the promising use of this tool for extensive screening.

## BACKGROUND

Since the outbreak of Corona Virus Disease 2019(COVID-19) in Wuhan, in December 2019, the epidemic has spread rapidly all over the world, which consequently brought great challenges to global public health(1). As of March 14, a total of 142,539 cases had been confirmed worldwide, with many more requiring tests(2). Currently, accurate nucleic acid detection plays a pivotal role in diagnosis and prevention of COVID-19, and reverse transcription polymerase chain reaction is the main method for nucleic acid detection(3). However, it is not feasible to use this time-and labor-consuming approach for screening a large growing number of suspected patients and asymptomatic infected patients(4). Rapid and universal screening method is essential to save medical resources, improve diagnosis efficiency and avoid cross-infection.

Taking advantage of the emerging machine learning technique, which enables analysis of the existing multidimensional data by applying appropriate algorithms for feature expression and classification, we have potential to improve the accuracy of diagnosis. For instance, in a recent study published in Nature, deep learning-based statistical model was used to improve breast cancer treatment and survival through identifying breast cancer patients with a high risk of long-term recurrence(5). In this study, we analyzed a variety of basic laboratory examinations indicators that related to host reactions to generate a free COVID-19 diagnosis aid APP. Then, the accuracy of this app was validated in an independent cohort(6).

## METHODS

### Patients and information collection

We have complied with all relevant ethical regulations for work with human subjects and the studies were approved by the Clinical Trials and Biomedical Ethics Committee of West China Hospital, Sichuan University. Through the West China hospital, a regional medical center, we collected the information of confirmed COVID-19 cases from various regional medical institutions, between 20th December 2019 and 10th February 2020. Diagnostic criteria were: RT-PCR of respiratory or blood samples was positive for SARS-CoV-2 (Severe Acute Respiratory Syndrome Coronavirus 2) nucleic acid; or viral gene sequencing of respiratory or blood sample was highly homologous with SARS-CoV-2. Controls were patients collected in the same site and the same time period, who were free of COVID-19 but had a diagnosis of viral pneumonia. Results of laboratory examinations on admission were collected which included age, gender and 35 indicators of whole blood count, coagulation test and biochemical examination. In addition, the independent validation cohort consisted of samples collected prospectively from one single medical center, using similar data collection strategy.

### Feature selection and model construction

Factors under consideration are age, sex and indicators of basic laboratory examinations(see Table 1). Using 70% samples from the derivation cohort (i.e., the training set), we first performed feature selection using Least Absolute Shrinkage and Selection Operator (LASSO) to select the valuable indicators of COVID-19. The main idea of LASSO for feature selection is to construct a first-order penalty function, which can shrink the regression coefficient (b) of each variable within a certain range, and eliminate the feature with a coefficient of 0, and finally obtain an optimally refined model(7). The penalty terms for LASSO are: sum (abs(b)) <= t. Then these indicators were used to construct a primitive model by multivariate logistic regression. The parameters were further adjusted through internal validation (i.e., the other 30% samples, the testing set) to obtain the optimized final model.

**Table 1.** Candidate of the model.

|  | Derivation cohort |  | Validation cohort |  |
| --- | --- | --- | --- | --- |
|  | Control | COVID-19 | Control | COVID-19 |
| SEX | female(27.41%) | female(31.07%) | female(31.21%) | female(30.56%) |
| AGE | 68.5(77,81) | 46(55,73) | 66(76,84) | 48(58,67) |
| PT | 11(11.75,13.25) | 10.7(11.8 ,13.4) | 11.1(12,13.7) | 11(11.7,12.7) |
| INR | 0.95(1.06,1.15) | 0.94(1.04,1.16) | 0.97(1.06,1.20) | 0.96(1.02,1.12) |
| APTT | 28.18(29.75,35.68) | 25.60(27.90,31.40) | 27.38(30.10,35.43) | 25.68(28.25,32.20) |
| TT | 17.10(18.00,19.13) | 16.40(17.20,18.10) | 17.20(18.20,19.60) | 16.98(17.90,19.10) |
| RBC | 3.13(3.76,4.13) | 3.66(4.25,4.76) | 3.10(3.68,4.15) | 3.47(3.98,4.44) |
| HGB | 93.25(110.50,126.50) | 111.00(129.00,148.00) | 91.75(110.00,123.00) | 103.00(119.00,135.00) |
| HCT | 0.29(0.35,0.39) | 0.35(0.38,0.44) | 0.29(0.34,0.38) | 0.32(0.36,0.40) |
| MCV | 88.13(92.20,96.30) | 88.50(91.50,95.00) | 88.90(93.25,97.20) | 88.40(91.65,95.53) |
| MCH | 29.00(29.90,31.20) | 28.90(30.70,32.10) | 28.90(30.00,31.30) | 29.10(0.40,31.70) |
| MCHC | 317.00(325.00,335.25) | 320.00(334.00,345.00) | 314.00(324.00,332.00) | 318.00(328.00,339.00) |
| RDW-SD | 44.58(47.45,51.13) | 41.90(44.50,49.80) | 45.08(49.20,54.10) | 42.65(46.20,50.25) |
| RDW-CV | 13.40(14.10,15.33) | 12.80(13.60,15.10) | 13.70(14.60,15.90) | 13.18(13.90,14.90) |
| PLT | 135.75(186.00,230.75) | 107.00(147.00,202.00) | 130.00(203.50,276.25) | 126.75(203.50,277.75) |
| WBC | 5.73(7.39,9.02) | 6.52(9.59,14.89) | 5.87(8.02,11.04) | 5.22(7.42,11.09) |
| TBIL | 6.20(9.75,15.18) | 8.50(12.20,17.50) | 6.90(9.65,14.48) | 7.60(10.30,14.40) |
| DBIL | 2.48(3.80,6.28) | 3.70(5.70,7.50) | 2.90(4.40,7.13) | 3.30(4.60,7.23) |
| IBIL | 3.78(4.95,6.98) | 4.00(6.10,9.10) | 3.30(4.60,7.10) | 3.40(5.10,7.50) |
| ALT | 12.00(19.00,30.25) | 18.00(27.00,55.00) | 13.00(21.00,37.00) | 19.00(31.50,57.00) |
| AST | 18.75(23.00, 36.00) | 22.00(36.00,68.00) | 19.00(25.50,39.25) | 21.00(33.00,55.00) |
| TP | 53.90(58.85,68.13) | 60.80(65.90,72.10) | 56.48(62.05,68.53) | 55.90(60.85,66.33) |
| ALB | 32.70(36.10,41.65) | 30.70 (34.10,39.00) | 31.30(34.75,38.53) | 29.90(32.95,37.23) |
| GLB | 19.43(23.80,28.45) | 27.50(31.80,35.90) | 22.50(26.50,30.93) | 23.38(27.00,31.60) |
| A/G | 1.33(1.51,1.91) | 0.90(1.11,1.32) | 1.11(1.35,1.61) | 1.03(1.22,1.46) |
| GLU | 4.92(6.00,7.64) | 6.31(7.33,9.67) | 5.49(7.20,9.40) | 5.10(6.24,8.40) |
| UREA | 4.60(6.05,8.25) | 4.20(6.20,10.40) | 4.58(6.69,9.43) | 3.40(4.73,7.00) |
| CREA | 50.50(60.50,103.50) | 60.00(75.00,94.00) | 50.00(65.50,90.00) | 51.00(61.00,77.00) |
| eGFR | 60.87(89.49,97.73) | 66.14(96.38,104.66) | 63.45(86.14,100.87) | 87.21(101.88,112.20) |
| Cys-C | 0.90(1.06,1.69) | 0.86(1.06,1.42) | 0.92(1.16,1.68) | 0.82(0.96,1.23) |
| URIC | 154.75(219.00,291.25) | 181.00(281.00,366.00) | 133.75(200.00,287.25) | 138.00(192.00,281.00) |
| TG | 0.71(0.93,1.23) | 0.98(1.17,1.80) | 0.76(1.03,1.34) | 0.94(1.30,1.82) |
| CK | 41.75(56.00,86.75) | 39.00(87.00,204.00) | 29.00(49.50,84.25) | 36.00(62.00,125.25) |
| NA | 135.30(139.20,141.83) | 131.40(135.50,139.10) | 135.95(139.70,142.60) | 135.80(139.15,141.70) |
| K | 3.72(4.08,4.38) | 3.32(3.76,4.09) | 3.73(4.08,4.51) | 3.57(3.96,4.38) |
| β-HBA | 0.07(0.12,0.23) | 0.08(0.23,0.38) | 0.07(0.11,0.22) | 0.06(0.12,0.30) |
| CA | 2.06(2.15,2.21) | 2.01(2.11,2.23) | 2.02(2.14,2.24) | 1.97(2.07,2.17) |

Data are presented as n (%) for categorical variables and as median (interquartile range) for continuous variables.
PT, prothrombin time; INR, International Normalized Ratio; APTT, activated partial thromboplastin time; TT,
thromboplastin time; RBC, red blood cell; HGB, hemoglobin; HCT, hematocrit; MCV, Mean Corpuscular Volume;

### Evaluation

In external validation cohort, we compared the predicted results, with the follow-up results to evaluate the diagnostic validity by positive predictive values, and negative predictive values. Finally, to facilitate further use in clinic setting, all model parameters and code were encapsulated as a visual application(APP), *COVID-19 Diagnosis Aid APP*.

### Statistical analysis

Continuous variables are represented by the median (upper and lower quartiles). Categorical variables are expressed in terms of frequency. LASSO algorithm was used for feature selection and the model was constructed by multivariate logistic regression. The diagnostic performance of the model was assessed by AUC. Calibration curves and Hosmer-Lemeshow test were used to evaluate the degree of overestimation or underestimation of the model. DCA were used to measure the net clinical benefits. The LASSO algorithm was performed by “glmmet” package. The logistic regression model was constructed by “glm” package. All statistical analyses were completed using R 3.5.0 version.

## RESULTS

### Participants and Clinical Characteristics

A total of 620 samples were included in the derivation cohort, among which 431 samples (211 COVID-19 vs. 220 Control) were in the training set and 189 samples (91 COVID-19 vs. 98 Control) were in testing set by simple randomization. The frequency of COVID-19 in the training set (48.96%) was not significantly different from that in the testing set (48.14%). We included 145 samples in the independent validation cohort.

### Model development and evaluation

Through Lasso regression screening and Multivariate logistic regression, 9 representative variables (age, Activated Partial Thromboplastin Time, Red Blood Cell Distribution Width-SD, Uric Acid, Triglyceride, Serum Potassium, Albumin/globulin, 3-Hydroxybutyrate, Serum Calcium) with good identification value were selected and constructed an optimized diagnostic model(see Figure 1). According to the suggestive information from the model, among 145 samples that used for validation, 80 samples were estimated as COVID-19 and 65 samples were considered COVID-19-free. Compared with the status determined by nucleic acid detection (the golden standard), we found 69 real COVID-19 samples were correctly identified by the app (69/80, positive predictive value =86.25%), while 55, out of the 65 COVID-19-free samples, were confirmed to be negative (55/65, negative predictive value =84.62%). The area under curve (AUC) of the model were 0.890 and 0.872 in the testing set and independent validation cohort, respectively(see Figure 2)(8). The calibration curve performed well and Hosmer-Lemeshow test results P value was much greater than 0.05(see Figure 3). The DCA quantitatively demonstrated high clinical net benefit over the entire probability threshold(see Figure 4)(9).

**Figure 1.**
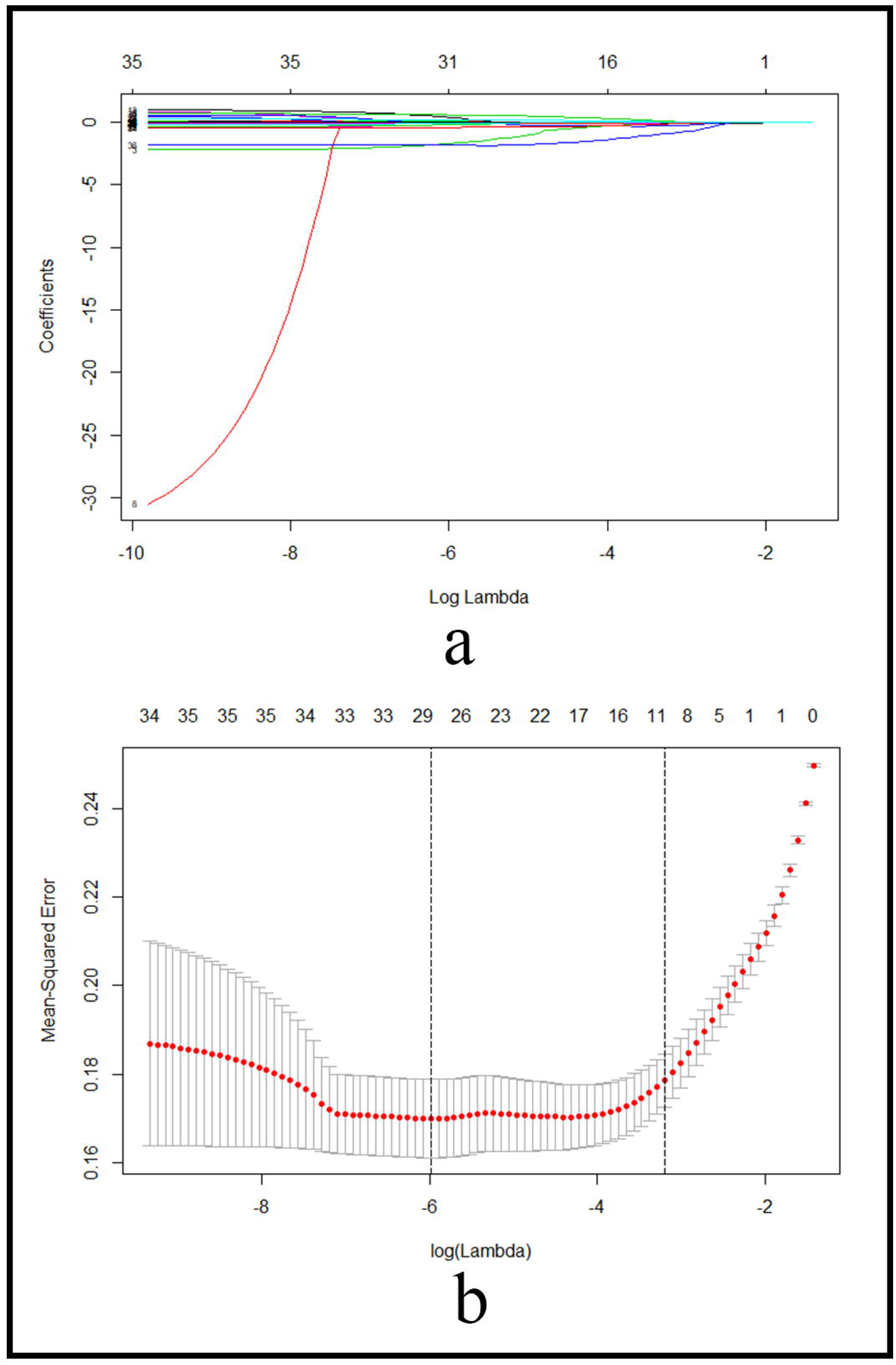
**(a) Coefficient diagram of LASSO variables.** Each curve in the figure represents the trajectory of the coefficient of an independent variable. The ordinate is the value of the coefficient. The lower abscissa, λ, is the parameter that controls the severity of the penalty. The upper abscissa is the number of non-zero coefficients in the model under the penalty parameter. **(b) Adjustment parameters in the LASSO model**. The lambda is screened by 10 folds cross-validation. A dashed vertical line is drawn at one standard error (1-SE standard) of the minimum and minimum standards. Lambda.1se corresponds to a model with good performance but the fewest number of arguments.

**Figure 2.**
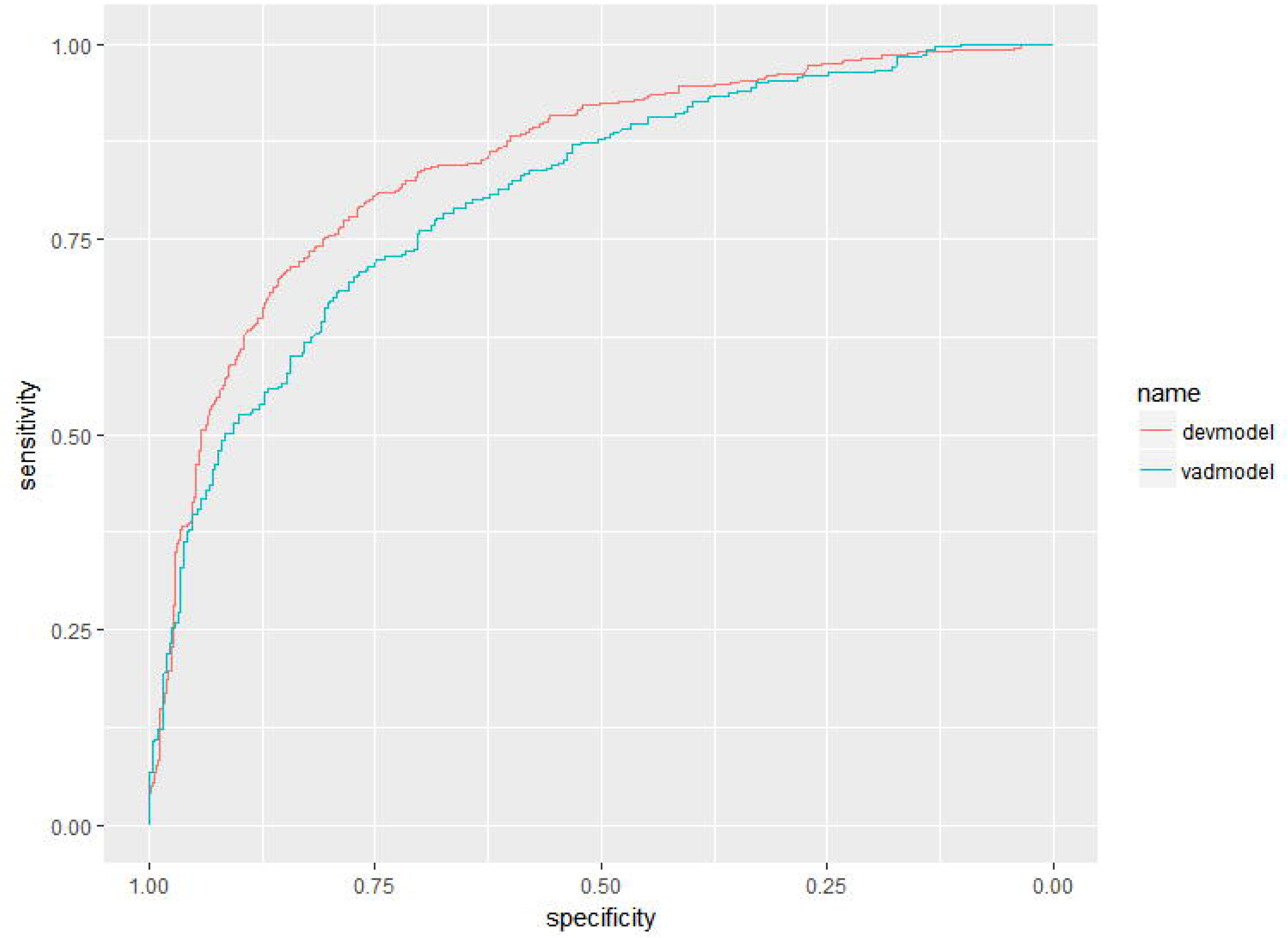
Receiver operator characteristic curve. The AUC were 0.890 and 0.872 in derivation and validation cohort respectively.

**Figure 3.**
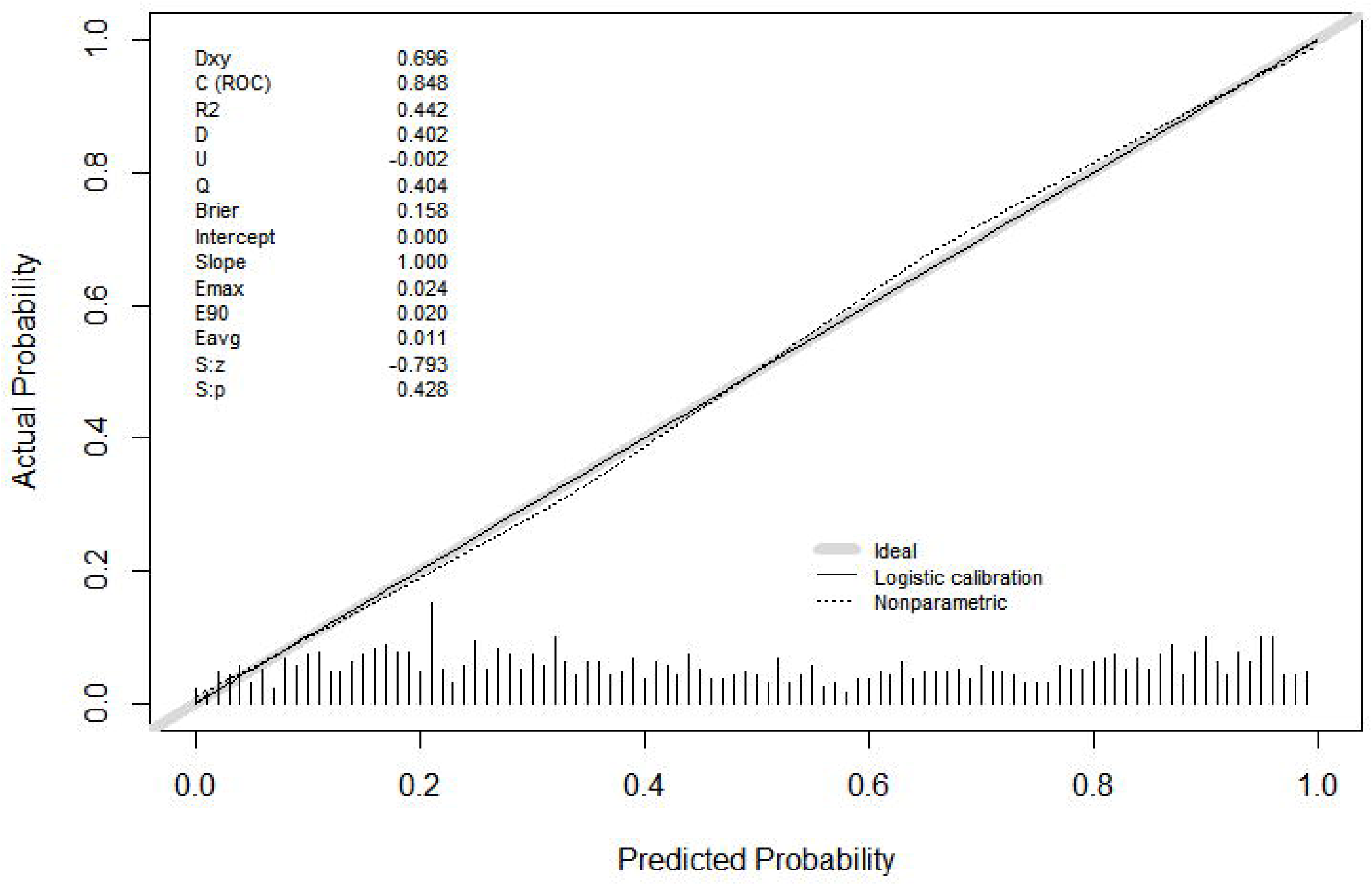
Calibration curves. The 45° shaded line represents the ideal prediction and the prediction probability is consistent with the actual observation probability. The Solid line represents the actual prediction of the model. The stapled histogram on the bottom line represents the distribution of patients’ predicted probability. Abbreviations: Dxy = Somer’s D rank correlation, R2 = Nagelkerke-Cox-Snell-Maddala-Magee R-squared index, D = Discrimination index, U = Unreliability index, Q = Quality index, Emax = maximum absolute difference in predicted and calibrated propabilities, S:z = Spiegelhalter Z-Test, S:p = two-tailed p-value of the Spiegelhalter Z-test.

**Figure 4.**
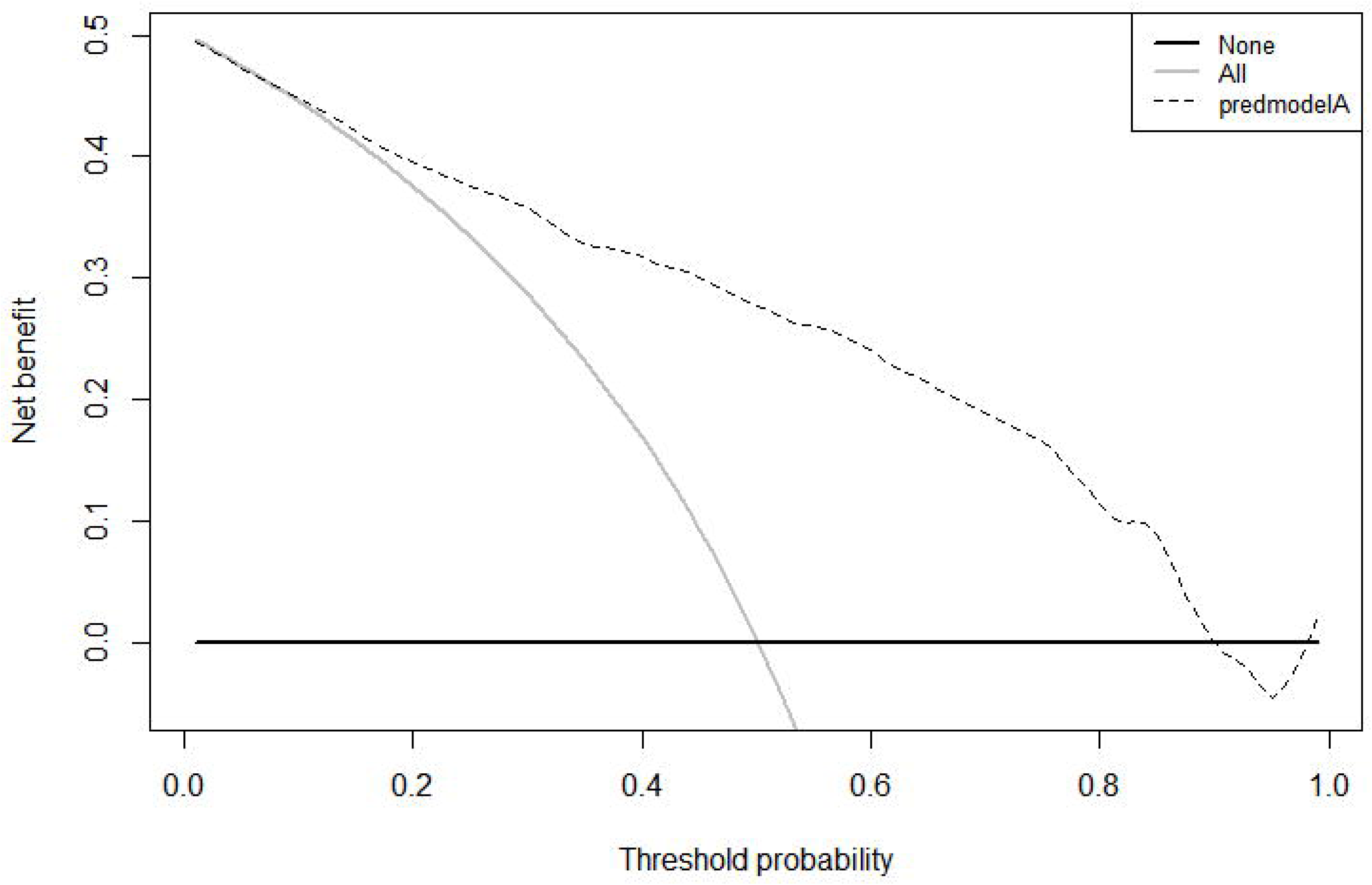
Decision curve analysis. The horizontal axis is the threshold probability of occurrence of COVID-19. The vertical axis shows the clinical benefits that patients may gain or lose using the APP. Dotted line: prediction model. Solid line: all patients were COVID-19. Horizontal line: all patients were not COVID-19.

### Construction of application

The model was encapsulated as an APP, *COVID-19 Diagnosis Aid APP*. The risk of COVID-19 can be obtained by inputting the required indicators into the *APP*. It will be online in the android store and IOS APP Store soon for facilitating the further application(see Figure 5).

**Figure 5.**
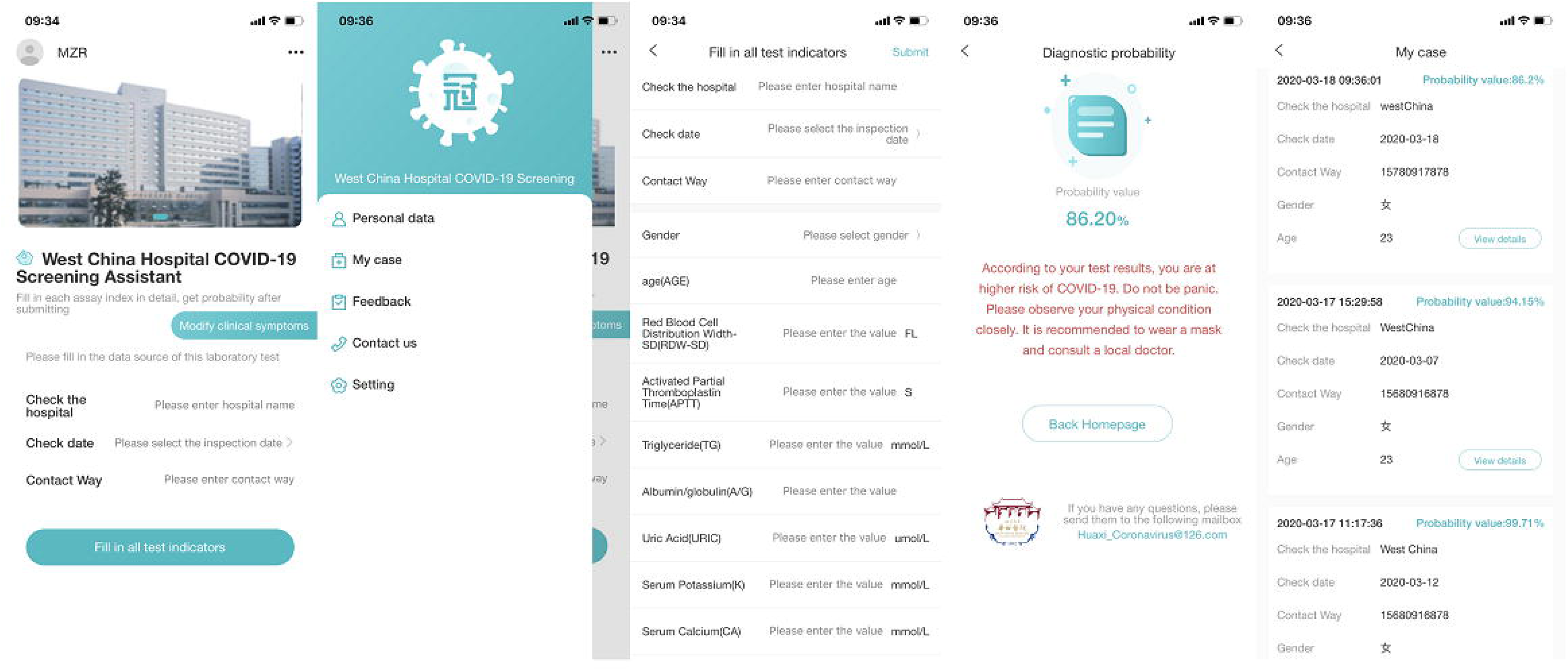
APP schematic. The model can show the probability of COVID-19 after inputing the corresponding index into the the APP.

## DISCUSSION

According to the latest epidemiological information of the World Health Organization(WHO), SARS-CoV-2 is spreading worldwide with an intensifying situation, and many countries including South Korea, Italy, and the United States have experienced outbreaks(10). Timely diagnosis, isolation and treatment are critical to control the global spread of COVID-19. However, nucleic acid testing is not always feasible or fully affordable in all regions. We therefore need more ways to realize the early and accurate community control and screening of suspicious people. To our knowledge, this is the first COVID-19 Diagnosis Aid APP based on laboratory tests for extensive screening. It has good accuracy and stability, which can help medical institutions to predict patients’ situation in advance and accurately invest the limited healthcare resources, so as to effectively control the spread of the epidemic.

Based on the optimized diagnostic model developed in current study, users can obtain the probability of COVID-19 according to the simple, easily available and reliable indicators from laboratory examinations. Given satisfiable model prediction performance (i.e., AUC was 0.872; positive predictive value and negative predictive value was reach 86.25% and 84.62%, respectively in independent validation cohort), the application of this app can provide reliable suggestion for further assessment of the index person, which has potential to reduce anxiety of public, as well as unnecessary hospital visit and nucleic acid testing.

It is undeniable that nucleic acid detection is one of the most reliable standards for the diagnosis of COVID-19, but we should also be aware of its shortcomings and deficiencies in practice. Nucleic acid testing requires professional technical platforms and physicians, and false negative results due to multiple reasons cannot be completely avoided(11). Instead, laboratory indicators that identified in our study are accessible, and can be widely used in community hospitals by family doctors to complete the early stage of suspected population screening.

On the other hand, based on the technological revolution brought by the Internet, this APP software installed on personal mobile can not only help to discover COVID-19 in the most convenient way, but also timely monitor dynamic trends of the prevalence of the contagious disease through data transmission to the Disease Control Center or professional medical institution(12, 13). This provides real-time data with predictive value, which can be reliable basis for more accurate health and epidemic prevention policies. Furthermore, this APP software is continuously upgradeable, with the possibility of extended scope of application. For example, we have added the data acquisition function which enabled the documentation of epidemiological and clinical features and a personalized reminder for necessary revisits. Also, with futher implementation of communication function, it is possible to establish a direct pre-connection between individuals and medical centers, which may benefit patients by avoiding unnecessary hospital visits or shorten waiting time spend before admission (14).

## CONCLUSION

*COVID-19 Diagnosis Aid APP* can efficiently and accurately calculate the infection probability through simple and easily-obtained laboratory examination results. It will promote the screening of a large number of suspected people, save limited medical resources, optimize the diagnosis process, and it can constantly learn, adapt and upgrade, which are vital to control the global spread of COVID-19.

## Data Availability

The data used to support the findings of this study are available from the corresponding author upon request.

## Ethics approval and consent to participate

The protocol of this study was approved by the West China Hospital, Sichuan University Medical Ethics Committee and conformed to the principles of the Declaration of Helsinki.

## The registry number for clinical trial

ChiCTR2000030542

## Consent for publication

Not applicable

## Data Availability Statement

The datasets used and analysed during the current study are available from the corresponding author on reasonable request.

## Authors’ contributions

Zirui Meng analyzed the data and was a major contributor in writing the manuscript. Minjin Wang designed the APP framework and was a major contributor in writing the manuscript. Huan Song analyzed the data and was a major contributor in writing the manuscript. Shuo Guo analyzed the data. Yanbing Zhou analyzed the data. Weimin Li analyzed patients’ medical records. Yongzhao Zhou analyzed patients’ medical records. Mengjiao Li collected the data. Xingbo Song verified the performance of the app. Yi Zhou verified the performance of the app. Qingfeng Li collected the data. Xiaojun Lu designed the functions of the app. Binwu Ying proposed research ideas and design research programs. All authors read and approved the final manuscript.

## Acknowledgements

The authors thank Beijing Zhongben Tech.co,.Ltd. for their technical contributions to this study.

